# Association Between Gastrointestinal Symptoms and Stroke Prevalence rate and Post-Stroke Mortality in Hypertensive Individuals: Evidence from a National Cohort

**DOI:** 10.64898/2025.12.01.25341395

**Authors:** Na Yin, Yebin Zou, Laicang Wang

## Abstract

**Background:** Stroke is a leading cause of mortality, and hypertension is a key risk factor. Gastrointestinal (GI) symptoms are frequent in stroke patients and may affect prevalence and prognosis via the brain-gut axis, but their role in hypertensive populations is unclear.

**Methods:** We analyzed data from 12,987 US adults in the National Health and Nutrition Examination Survey (NHANES) (2005–2010). Stroke history was self-reported, and GI symptoms (chronic constipation or diarrhoea) were categorized using the Bristol Stool Scale. Weighted multivariable logistic regression was employed to evaluate the relationship between GI symptoms and stroke in individuals with hypertension, while Cox models were utilized to examine their influence on cardiovascular and all-cause mortality in hypertensive stroke survivors, fully adjusting for covariates.

**Results:** Among hypertensive individuals, diarrhea (OR = 1.65, 95% CI: 1.08–2.54) and overall gastrointestinal symptoms (OR = 1.68, 95% CI: 1.20–2.35) were significantly associated with an increased prevalence rate of stroke after full adjustment. In the overall population, diarrhea (OR = 1.60, 95% CI: 1.00–2.53) and gastrointestinal symptoms (OR = 1.55, 95% CI: 1.12–2.13) also showed significant associations with stroke. In hypertensive stroke survivors, constipation was associated with an increased risk of cardiovascular mortality (HR = 2.15, 95% CI: 1.07–4.31).

**Conclusions:** In this study, gastrointestinal symptoms were found to be associated with stroke prevalence in hypertensive adults. Notably, constipation was prospectively linked to an increased risk of cardiovascular mortality in hypertensive stroke survivors. These findings suggest a gut-brain interaction and highlight gut health’s importance for prognosis in high-risk populations.

## 1. Introduction

Stroke is one of the leading causes of death and disability worldwide, with high incidence and mortality rates. According to the World Health Organization, it is the second most common cause of death globally, resulting in millions of deaths each year and placing a significant strain on healthcare systems^1^. Not only does stroke severely impact patients’ quality of life, it may also lead to long-term functional impairments and economic losses. Studies have shown that stroke patients often experience various complications, with gastrointestinal symptoms such as constipation, diarrhoea and gastrointestinal bleeding being particularly common and further worsening clinical outcomes^2,3^. Therefore, improving our understanding of the epidemiological features of stroke and its associated complications is crucial for enhancing clinical management and preventive strategies.

Hypertension is one of the most significant modifiable prevalence rates for stroke. Uncontrolled long-term hypertension markedly increases the risk of both ischemic and haemorrhagic stroke^4,5^. Epidemiological studies indicate that the incidence of stroke in hypertensive individuals is several times higher than in populations with normal blood pressure, and that the severity of hypertension is associated with an increased risk of stroke^4^. Furthermore, hypertension can worsen stroke pathology by promoting atherosclerosis, vascular endothelial dysfunction and cerebral haemodynamic abnormalities^5,6^. It is also important to note that hypertensive patients often present with comorbid metabolic disorders, such as diabetes and chronic kidney disease, which may synergistically elevate stroke risk^7,8^. Therefore, in hypertensive populations, identifying and managing additional prevalence rates (e.g. gastrointestinal symptoms) could help to reduce the incidence of stroke and its adverse outcomes.

There is growing evidence to suggest that gastrointestinal symptoms (e.g. constipation and diarrhoea) may be closely linked to the occurrence and progression of stroke^9^. Research indicates that gastrointestinal dysfunction (e.g. gastric disorders, functional bowel diseases and infectious intestinal diseases) can independently increase the prevalence rate of a future ischemic stroke, even when conventional vascular prevalence rates are taken into account^9^. Furthermore, patients who have had a stroke frequently develop gastrointestinal complications, such as constipation and bleeding. These complications hinder recovery and may also exacerbate neurological damage via the ‘brain-gut axis’ mechanism^1–3^. In populations with hypertension, the prevalence of gastrointestinal symptoms and their interaction with cardiovascular and metabolic disturbances may further increase the risk of stroke and cardiovascular mortality^10^. Therefore, investigating the relationship between gastrointestinal symptoms and stroke/cardiovascular mortality in hypertensive patients could reveal new targets for clinical intervention.

In summary, while the high incidence of stroke is a significant global public health concern, research into the relationship between gastrointestinal symptoms (e.g. constipation and diarrhoea) and stroke prevalence rate in hypertensive populations is limited. Current evidence indicates that hypertension is a significant stroke prevalence rate and that gastrointestinal symptoms are common among stroke patients. This study is based on two primary hypotheses. Firstly, there is a positive correlation between gastrointestinal symptoms and stroke prevalence among hypertensive individuals. Secondly, these symptoms are associated with an increased risk of mortality among hypertensive stroke survivors. Data were obtained from the National Health and Nutrition Examination Survey (NHANES) database, which included 12,987 Americans between 2005 and 2010.

## 2. Methods

### 2.1. Study population

The National Health and Nutrition Examination Survey (NHANES), initiated by the National Center for Health Statistics (NCHS), constitutes a nationally representative, cross-sectional assessment of the health and nutritional status among non-institutionalized U.S. residents. Reliable population-level estimates are generated through the application of multiple investigative methods, encompassing structured interviews, clinical examinations, and laboratory analyses. The sampling strategy is built upon a multistage probability design with stratification to secure demographic representativeness. Ethical endorsement for the study was provided by the NCHS Review Board, with written informed consent obtained from all enrollees. Comprehensive documentation on the operational methodology and recruitment protocols is publicly accessible online (http://www.cdc.gov/nchs/nhanes).

This study used data from three consecutive rounds of the National Health and Nutrition Examination Survey (NHANES), which were conducted between 2005 and 2010. Initially, 31,034 participants were enrolled. Of these, 16,448 were excluded due to missing data on stroke, gastrointestinal disease or hypertension, and a further 1,599 were excluded due to missing covariate data. The final analysis cohort comprised 12,987 hypertensive individuals and was used to investigate the association between gastrointestinal symptoms and stroke prevalence. When subsequently analysing mortality among hypertensive stroke survivors, we excluded an additional eight participants with missing mortality data, resulting in a final sub-cohort of 473 individuals. The participant selection process is detailed in **Figure 1**.

**Fig. 1.**
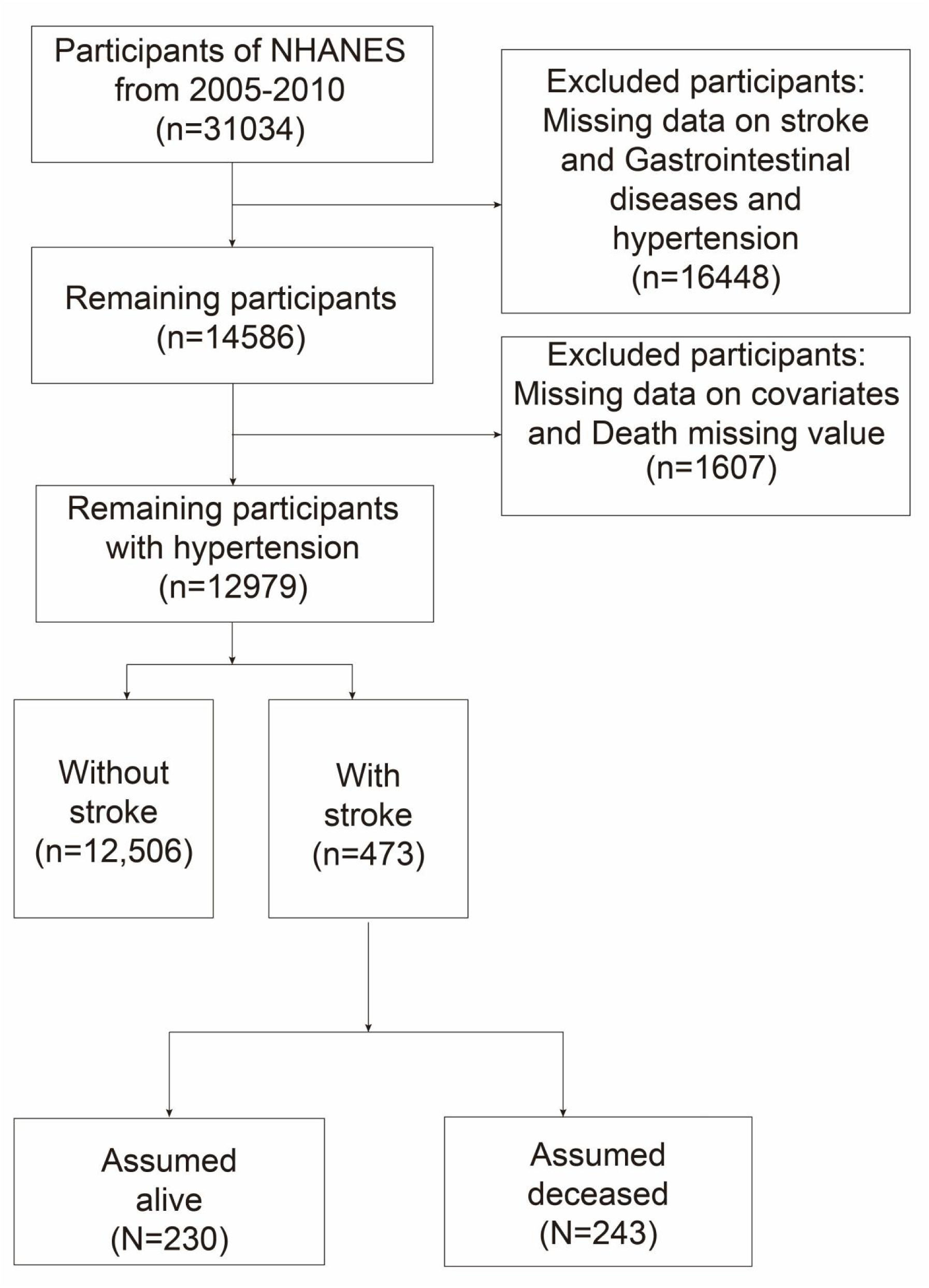

### 2.2. Measurement of stroke

Participants’ stroke history was assessed using medical history data collected through the NHANES survey. During the survey, trained healthcare personnel asked, “Have you ever been diagnosed with a stroke by a doctor or other healthcare professional?” Those who responded positively to this question were classified as having had a previous stroke^11^.

### 2.3. Measurement of Chronic diarrhea and constipation

Participants were categorized according to their typical stool patterns based on the Bristol Stool Scale. During the NHANES examination, participants were shown a card illustrating the Bristol Stool Form Scale and asked to indicate the number that best matched their usual or most common type of stool. In line with previous studies, chronic constipation was defined as consistently having type 1 (hard lumps) or type 2 (lumpy, sausage-shaped) stools. Conversely, chronic diarrhoea was characterized by types 6 (fluffy, mushy fragments with ragged edges) or 7 (entirely liquid, watery stools with no solid material). Participants reporting stool types falling outside these categories were considered to have normal bowel function^12^.

### 2.4. Measurement of hypertension

Blood pressure measurements were performed in accordance with the standardized protocols established by the American Heart Association. Following three consecutive BP measurements taken under resting conditions, mean systolic blood pressure (SBP) and diastolic blood pressure (DBP) values were computed. According to the 2017 American Heart Association/American College of Cardiology (AHA/ACC) guideline, hypertension was defined as SBP ≥130 mmHg and/or DBP ≥80 mmHg. Additionally, participants who responded affirmatively to the question regarding a prior diagnosis of high blood pressure were also categorized as having hypertension^13^.

### 2.5. Covariates

The analysis accounted for a set of pre-specified covariates encompassing Socio-demographic factors (sex, age, race, marital status, educational attainment, body mass index [BMI] and poverty-to-income ratio [PIR]), behavioral factors (alcohol consumption and smoking status) and clinical comorbidities (diabetes and hyperlipidemia).

Smoking status was defined based on self-reported lifetime cigarette consumption and current use. Participants who reported having smoked at least 100 cigarettes and were currently smoking were classified as current smokers. Those who had smoked more than 100 cigarettes but had quit were considered former smokers, while individuals with a lifetime consumption of fewer than 100 cigarettes were deemed non-smokers. Alcohol intake was categorized as none/moderate (women: 0–1, men: 0–2 drinks/day), heavy (women: 2–3, men: 3–4 drinks/day), or binge drinking (women: ≥4, men: ≥5 drinks/day)^14^. Diabetes mellitus was defined as a physician-confirmed diagnosis, a fasting plasma glucose level ≥126 mg/dL, or a glycated hemoglobin (HbA1c) value of 6.5% or higher. Hyperlipidemia was defined per NCEP ATP III or ACC/AHA guidelines, meeting any of the following criteria: total cholesterol ≥200 mg/dL, LDL-C ≥130 mg/dL, HDL-C <40 mg/dL (men) or <50 mg/dL (women), triglycerides ≥150 mg/dL, current use of lipid-lowering medication, or a self-reported physician diagnosis^15^.

### 2.6. Statistical analysis

Categorical variables are described using weighted n (%) and continuous variables are represented using mean and standard error (SE). Differences between groups were assessed using the chi-square test (categorical variables) and weighted t-test (continuous variables) or Mann–Whitney U test (continuous non-normally distributed variables), respectively. Weighted logistic regression models were used to calculate the adjusted odds ratio (OR) and 95% confidence interval (CI) for the association between gastrointestinal health and stroke in individuals with hypertension. The hazard ratio (HR) and its 95% confidence interval (CI) were estimated using the weighted Cox proportional hazards model to assess the impact of gastrointestinal health on mortality risk in individuals with hypertension and stroke. Follow-up duration served as the baseline time metric. Three models were employed for covariate adjustment: Model 1 was unadjusted, Model 2 was adjusted for age, sex and ethnicity, and Model 3 was adjusted for all the covariates in Model 2, plus educational attainment, marital status, BMI, PIR, alcohol consumption, smoking status, diabetes and hyperlipidaemia. A two-sided p-value of <0.05 was deemed to be statistically significant. All statistical analyses were conducted using the software EmpowerStats and the R statistical software packages.

## 3. Results

### 3.1. Baseline characteristics

The baseline characteristics of the study participants are summarized in **Table 1**. The study included individuals with a mean age of 56.51 years, with the cohort distributed almost equally by sex (49.83% male; 50.17% female). Significant differences were observed in several demographic and clinical variables between groups.Notably, the prevalence of gastrointestinal symptoms was 14.24% in the total population, with a notably higher proportion observed in the stroke group (24.08%, *P* < 0.0001). Similarly, diarrhoea was reported by 7.92% of the total population, but its prevalence was markedly higher among stroke participants (13.73%, *P* = 0.0016). Constipation was present in 6.32% of the total sample, with a significantly higher rate observed in the stroke group (10.35%, *P* = 0.0156).Other characteristics also showed significant differences, including age, poverty-income ratio (PIR), educational attainment, marital status, smoking and alcohol consumption, and diabetes status (all *P* < 0.05).

**Table 1.**
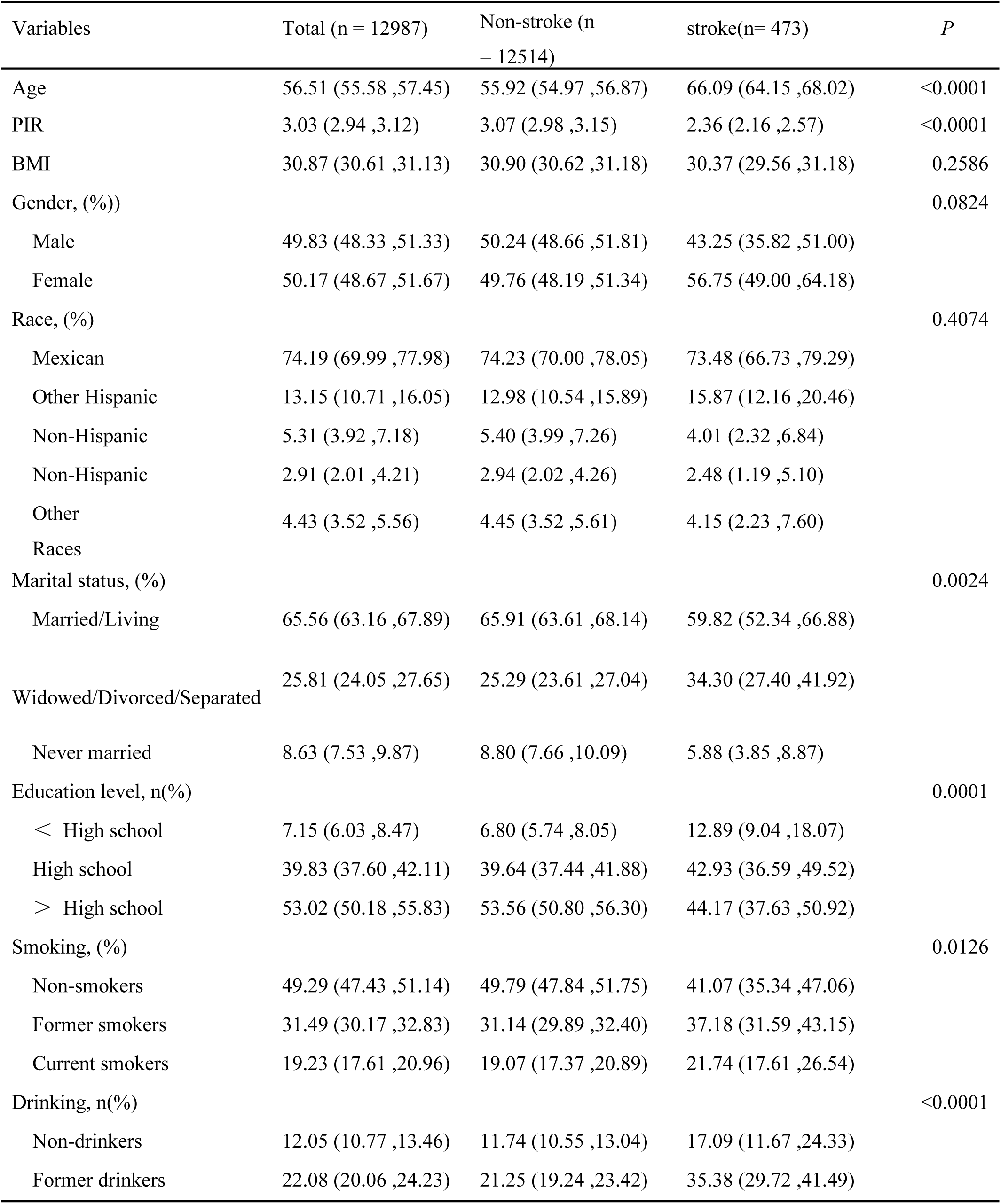
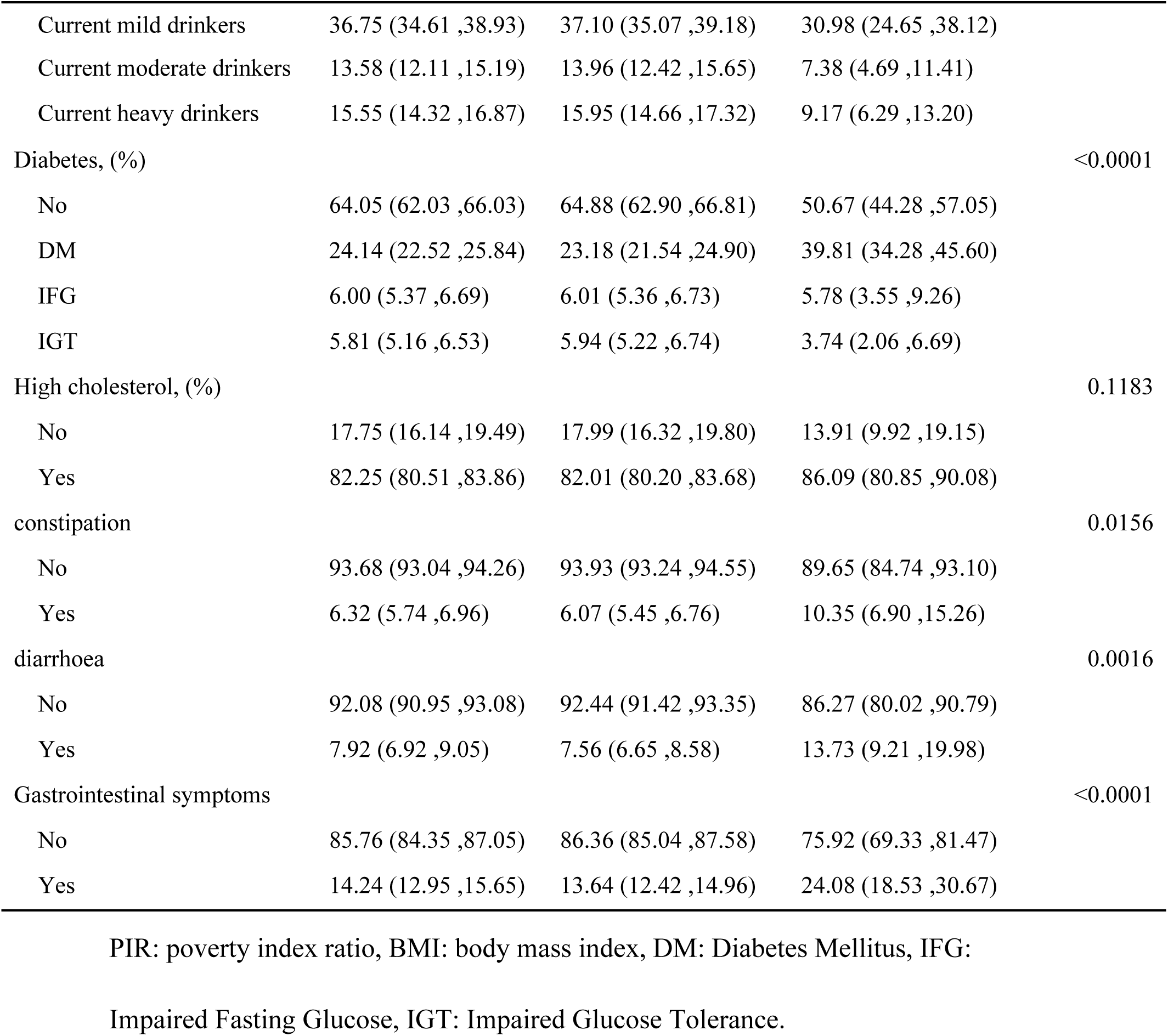
Baseline characteristics of participants.

### 3.2. Association between between Gastrointestinal Health and stroke in Individuals with hypertension

**Table 2** shows the associations between gastrointestinal symptoms and stroke, categorized by hypertension status and the entire population, as determined by multivariable logistic regression models. After full adjustment for age, sex, race, education, body mass index, family income-to-poverty ratio, marital status, alcohol consumption, smoking status, diabetes and hyperlipidaemia (Model 3), several significant associations were observed. Among hypertensive patients, diarrhea (odds ratio = 1.65, 95% confidence interval: 1.08–2.54, *P*=0.030) and the presence of overall gastrointestinal symptoms (odds ratio = 1.68, 95% confidence interval: 1.20–2.35, *P*=0.0059) were significantly associated with increased stroke incidence. Specifically, compared to those without diarrhea, individuals with diarrhea had a 65% higher stroke incidence rate. Similarly, those with gastrointestinal symptoms had a 68% higher incidence rate than those without symptoms. In the fully adjusted model for the overall population, diarrhea (OR=1.60, 95% CI: 1.00–2.53, *P*=0.0505) and gastrointestinal symptoms (OR=1.55, 95% CI: 1.12–2.13, *P*=0.0138) remained significantly associated with stroke, increasing the stroke prevalence by 60% and 55%, respectively, compared to those without these symptoms.

**Table 2.** The relationships between Gastrointestinal Health and stroke.

| Variables | Model1 |  | Model2 |  | Model3 |  |
| --- | --- | --- | --- | --- | --- | --- |
|  | OR (95% CI) | *P* value | OR (95% CI) | *P* value | OR (95% CI) | *P* value |
| **Relationships between Gastrointestinal Health and stroke without hypertension** |  |  |  |  |  |  |
| constipation | 1.01 (0.44, 2.31) | 0.9770 | 1.00 (0.43, 2.34) | 0.6646 | 0.93 (0.41, 2.15) | 0.8732 |
| diarrhea | 2.00 (0.80, 5.03) | 0.1454 | 1.63 (0.64, 4.14) | 0.3128 | 1.39 (0.54, 3.62) | 0.5012 |
| Gastrointestinal symptoms | 1.50 (0.74, 3.03) | 0.2681 | 1.34 (0.65, 2.74) | 0.4306 | 1.18 (0.57, 2.45) | 0.6517 |
| **Relationships between Gastrointestinal Health and stroke with hypertension** |  |  |  |  |  |  |
| constipation | 1.79 (1.11, 2.88) | 0.0212 | 1.60 (0.97, 2.63) | 0.0705 | 1.49 (0.85, 2.61) | 0.1786 |
| diarrhea | 1.95 (1.28, 2.96) | 0.0033 | 1.90 (1.22, 2.96) | 0.0070 | 1.65 (1.08, 2.54) | 0.0300 |
| Gastrointestinal symptoms | 2.01(1.45, 2.78) | 0.0001 | 1.89 (1.37, 2.61) | 0.0004 | 1.68 (1.20, 2.35) | 0.0059 |
| **Relationships between Gastrointestinal Health and stroke** |  |  |  |  |  |  |
| constipation | 1.59 (1.05,2.41) | 0.0349 | 1.45 (0.92, 2.27) | 0.1143 | 1.34 (0.81, 2.21) | 0.2622 |
| diarrhea | 1.96 (1.28, 3.00) | 0.0036 | 1.85 (1.19, 2.88) | 0.0096 | 1.60 (1.00, 2.53) | 0.0505 |
| Gastrointestinal symptoms | 1.89 (1.41, 2.54) | 0.0001 | 1.76 (1.29, 2.39) | 0.0009 | 1.55 (1.12, 2.13) | 0.0138 |

OR: Odds Ratio, CI: Confidence Interval.
Model1: Crude.
Model2: Adjust: Gender, Race, Age.
Model3: Adjust: Gender, Race, Age, Education level, Marital status, Smoking,
Drinking, PIR, BMI, Diabetes, High cholesterol.

### 3.3. Association between Gastrointestinal Health Indicators and Mortality in Hypertensive Stroke Survivors

**Table 3** details the association between gastrointestinal health status and mortality risk among hypertensive stroke survivors. In the fully adjusted model (Model 3), which controlled for demographic, socioeconomic, and clinical confounders, individuals with constipation exhibited a significantly elevated risk of cardiovascular disease mortality. The hazard ratio (HR) reached 2.15 (95% confidence interval 1.07–4.31, *P*=0.032), representing a 115% increased risk compared to those without constipation. For all-cause mortality, a trend toward increased risk was observed despite weaker and non-statistically significant associations (HR 1.39, 95% CI 0.88–2.21, *P* = 0.161).Conversely, in Model 3, the presence of diarrhea was not associated with a significant increase in the risk of all-cause mortality (HR 0.87, 95% CI 0.55–1.37, *P* = 0.551) or cardiovascular disease mortality (HR 0.51, 95% CI 0.21–1.24, *P* = 0.137). Similarly, after full adjustment, the composite indicator of gastrointestinal symptoms showed no significant association with all-cause mortality (HR 1.09, 95% CI 0.77–1.56, *P* = 0.616) or cardiovascular disease mortality (HR 1.09, 95% CI 0.60–1.97, *P* = 0.773).

**Table 3.**
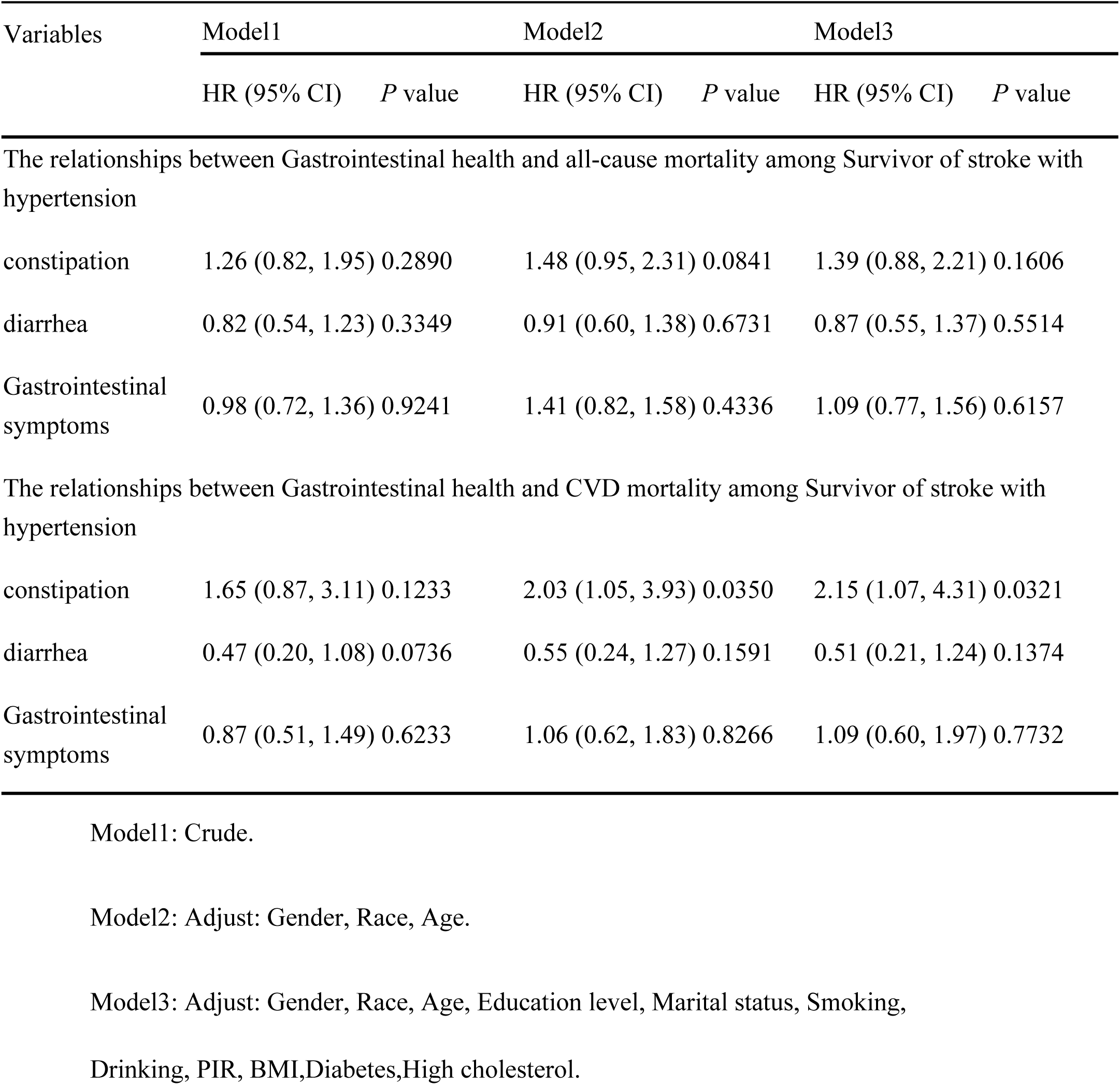
The relationships between Gastrointestinal health and mortality among Survivor of stroke with hypertension.

## 4. Discussion

Using a cross-sectional analysis of participants in the NHANES cohort, a significant association was identified between gastrointestinal symptoms and stroke among individuals with hypertension. This association remained consistent following adjustment for demographic, socioeconomic and clinical covariates. Specifically, diarrhoea and overall gastrointestinal symptoms were strongly associated with an increased prevalence rate of stroke in this population. Furthermore, constipation was significantly associated with an elevated risk of cardiovascular disease mortality among hypertensive stroke survivors, highlighting its potential prognostic importance.

Our findings resonate with, yet significantly build upon, the recent work of Du et al. (2023). While both studies use the NHANES database and confirm a link between gastrointestinal dysfunction and stroke, important methodological and conceptual differences highlight the novel contributions of our analysis. Firstly, whereas Du et al. examined the general adult population, our study specifically targeted hypertensive individuals — a high-risk subgroup in whom the gut–stroke interaction may be particularly significant. Secondly, we expanded the range of gastrointestinal symptoms to encompass not only constipation, but also diarrhoea and a composite GI symptom score. This revealed that both constipation and diarrhoea are independently associated with stroke prevalence in this population. Most importantly, and representing a fundamental advance beyond the prior study, we evaluated post-stroke mortality. Our identification of constipation as a significant predictor of cardiovascular mortality in hypertensive stroke survivors adds a crucial prognostic dimension that goes beyond the previously reported cross-sectional associations. The differences in population focus, symptom spectrum and outcome measures illustrate the complementary yet distinct contributions of the two studies to our understanding of the gut–stroke axis^16^.

When compared with the systematic review and meta-analysis by Li et al. (2017), which focused on constipation incidence among stroke patients, our findings further underscore the clinical relevance of gastrointestinal symptoms in stroke. However, key differences emerge in both focus and methodology. Li et al. examined the incidence of constipation in patients who had already experienced a stroke, reporting a pooled constipation incidence of 48%, with higher rates in hemorrhagic stroke and during rehabilitation. In contrast, our study investigated the association between gastrointestinal symptoms (constipation and diarrhea) and stroke prevalence rate in a hypertensive population, as well as their impact on post-stroke mortality. Methodologically, our analysis used cross-sectional and survival data from NHANES, whereas Li et al. synthesized evidence from eight previous studies. Notably, our results identified constipation as a predictor of cardiovascular mortality in hypertensive stroke survivors and diarrhea as a prevalence rate for stroke in this population. These distinctions reflect differing research questions: our study emphasizes hypertension as a precursor and modifier of stroke prevalence rate, while Li et al. focused on constipation prevalence after stroke^17^.

A comparison with the recent study by Ananth et al. (2022), which analyzed U.S. stroke mortality trends from 1975 to 2019, reveals both parallels and divergences. Both studies utilized large-scale U.S. population data and applied rigorous statistical methods to derive public health insights related to stroke. However, the studies differ substantially in scope and design. Ananth et al. performed a temporal trend analysis of stroke mortality by subtype using national death records, whereas our study adopted a cross-sectional approach with NHANES data to examine the association between gastrointestinal symptoms and stroke prevalence rate—as well as post-stroke mortality—among hypertensive individuals. Importantly, our findings introduce gastrointestinal dysfunction as a novel prevalence rate in stroke etiology and prognosis, a dimension not explored in the earlier mortality trend analysis. These differences underscore the complementary value of the two studies: one tracking long-term epidemiological trends, and the other identifying modifiable prevalence rates within a high-risk subgroup, together advancing the understanding of stroke prevention and management^18^.

This study revealed a positive correlation between diarrhoea and stroke prevalence rate in populations with hypertension, as well as a positive association between constipation and mortality in survivors of hypertensive stroke, which may be explained by several Pathophysiological mechanisms. Firstly, with regard to the association between diarrhoea and stroke, the literature suggests that gut microbiota dysbiosis may be a key mediating factor ^19,20^. The chronic inflammatory state in hypertensive patients may be exacerbated by diarrhoea-induced impairment of intestinal barrier function. This allows bacterial metabolites and inflammatory factors to enter the circulatory system via the ‘gut-brain axis’, thereby aggravating cerebrovascular endothelial dysfunction and atherosclerosis progression^19,21^. In particular, immune cells that are overactivated after a stroke can cause severe intestinal dysfunction^21^, and diarrhoea may reflect the severity of this gut microbial imbalance^22^. Secondly, consumption of highly processed foods is associated with an increased risk of both constipation and diarrhoea in hypertensive patients^23^, which may further exacerbate the vicious cycle between gut microbiota disruption and cerebrovascular diseases.

Concerning the association between constipation and mortality in stroke survivors, potential explanations include: 1) constipation as a marker of autonomic dysfunction reflecting more severe neurological damage^24^;2) gut microbiota alterations caused by chronic constipation exacerbating neuroinflammation through the microbiota-gut-brain axis and affecting post-stroke recovery^25^;3) certain medications commonly used by patients with constipation having adverse cardiovascular effects^24^. Studies have shown that individuals with constipation have a significantly higher risk of neurological disorders (stroke HR=1.35, dementia HR=1.50 and Parkinson’s disease HR=1.56)^24^, which may explain the increased mortality observed in stroke survivors. Furthermore, post-stroke intestinal dysfunction is closely related to a poor prognosis^26^, and constipation may be a clinical manifestation of this dysfunction. Notably, interventions targeting brain-gut interactions may improve outcomes in ischemic stroke^26^, providing potential avenues for future research.

The present study is subject to several limitations. Firstly, the cross-sectional design of the NHANES database means that causality between gastrointestinal symptoms and stroke or mortality cannot be inferred. Secondly, despite adjusting for a wide range of confounders, residual confounding from unmeasured factors such as medication use, detailed dietary habits or physical activity levels cannot be entirely excluded. Thirdly, the assessment of gastrointestinal symptoms relied on self-reported stool patterns based on the Bristol Stool Scale, which may be subject to recall bias or misclassification. Finally, as the study population consisted of US adults, our findings may not be generalisable to other ethnic or geographic populations.

Despite these limitations, our study has several notable strengths. To our knowledge, this is one of the first studies to examine the joint association between a range of gastrointestinal symptoms (including constipation and diarrhoea) and stroke prevalence rate in a population at high risk of hypertension, as well as the longitudinal association with cardiovascular mortality in hypertensive stroke survivors. Using a large, nationally representative sample (NHANES) enhances the generalisability and statistical power of our findings. Furthermore, we employed a comprehensive analytical approach, utilizing both multivariable logistic regression and Cox proportional hazards models to assess different aspects of the relationship. Our findings remained robust when accounting for a wide array of potential confounders, including socio-demographic, behavioral, and clinical comorbidities, thereby strengthening the validity of our conclusions.

## 5. Conclusion

In this national cohort, gastrointestinal symptoms, particularly diarrhea, were associated with a higher stroke prevalence among hypertensive individuals. Crucially, in prospective follow-up, constipation was independently associated with increased cardiovascular mortality in hypertensive stroke survivors. Although the mortality association implies temporality, future prospective studies are needed to establish causality and elucidate the underlying mechanisms of the brain-gut axis in cerebrovascular disease.

## Data Availability

All relevant data are contained in the manuscript and its supporting information files.

https://wwwn.cdc.gov/nchs/nhanes/

## Statement on the use of AI tools

No AI tools were used in manuscript preparation or data analysis

## Ethics statement

In accordance with local legislation and institutional requirements, ethical approval was not necessary for this study. Similarly, written informed consent was not required from participants or their legal guardians under national regulations.

## Publisher’s note

All statements made in this article are solely those of the authors and do not necessarily reflect those of their affiliated organizations, the publisher, the editors, or the reviewers. Any products evaluated or claims made by manufacturers are not guaranteed or endorsed by the publisher.

## CRediT authorship contribution statement

Na Yin: Writing – original draft, Formal analysis, Data curation, Conceptualization.

Yebin Zou: Writing – review & editing, Methodology, Investigation.

Laicang Wang: Writing – review & editing, Supervision, Project administration, Funding acquisition.

## Acknowledgments

We sincerely appreciate all participants who contributed to this study.

## References

1 Ye, D. et al. Exploratory Investigation of Intestinal Structure and Function after Stroke in Mice. Mediators of inflammation 2021, 1315797, doi:10.1155/2021/1315797 (2021).

2 Tan, B. Y. Q., Paliwal, P. R. & Sharma, V. K. Gut Microbiota and Stroke. Annals of Indian Academy of Neurology 23, 155–158, doi:10.4103/aian.AIAN_483_19 (2020).

3 Yong, H. Y. F., Ganesh, A. & Camara-Lemarroy, C. Gastrointestinal Dysfunction in Stroke. Seminars in neurology 43, 609–625, doi:10.1055/s-0043-1771470 (2023).

4 Hu, J., Zheng, X., Shi, G. & Guo, L. Associations of multiple chronic disease and depressive symptoms with incident stroke among Chinese middle-aged and elderly adults: a nationwide population-based cohort study. BMC geriatrics 22, 660, doi:10.1186/s12877-022-03329-4 (2022).

5 Dan, X. et al. Differential Analysis of Hypertension-Associated Intestinal Microbiota. International journal of medical sciences 16, 872–881, doi:10.7150/ijms.29322 (2019).

6 Jaroonpipatkul, C. et al. Depressive symptoms due to stroke are strongly predicted by the volume and location of the cerebral infarction, white matter hyperintensities, hypertension, and age: A precision nomothetic psychiatry analysis. Journal of affective disorders 309, 141–150, doi:10.1016/j.jad.2022.04.041 (2022).

7 Sarfo, F. S. et al. Administration of a pictorial questionnaire to screen for stroke among patients with hypertension or diabetes in rural Ghana. Journal of the neurological sciences 373, 289–294, doi:10.1016/j.jns.2017.01.022 (2017).

8 Kashiwagi, Y., Okuno, H., Nishida, S., Ishii, H. & Yamanaka, G. A case of chronic kidney disease with refractory periodic vomiting and hypertension in a pediatric patient. CEN case reports 14, 103–107, doi:10.1007/s13730-024-00905-y (2025).

9 Roth, W. H. et al. Gastrointestinal Disorders and Risk of First-Ever Ischemic Stroke. Stroke 51, 3577–3583, doi:10.1161/strokeaha.120.030643 (2020).

10 Dawoodi, S., Dawoodi, I. & Dixit, P. Gastrointestinal problem among Indian adults: Evidence from longitudinal aging study in India 2017-18. Frontiers in public health 10, 911354, doi:10.3389/fpubh.2022.911354 (2022).

11 Fan, X. et al. The relationship between CALLY index and stroke in hypertensive patients: insights from NHANES. Frontiers in nutrition 12, 1592641, doi:10.3389/fnut.2025.1592641 (2025).

12 Zhuang, Y., Li, L., Sun, J., Zhang, Y. & Dai, F. Association of body roundness index with chronic diarrhea and constipation, NHANES 2005-2010. Journal of health, population, and nutrition 44, 50, doi:10.1186/s41043-025-00793-7 (2025).

13 Yuan, M. et al. Hypertension and NAFLD risk: Insights from the NHANES 2017-2018 and Mendelian randomization analyses. Chinese medical journal 137, 457–464, doi:10.1097/cm9.0000000000002753 (2024).

14 Lai, S. et al. Association Between Dietary Fiber Intake and Stroke Among US Adults: From NHANES and Mendelian Randomization Analysis. Stroke 56, 1786–1798, doi:10.1161/strokeaha.124.049093 (2025).

15 Ma, S., Miao, Y. & Wu, X. Fermented dairy products intake and stroke risk: analyses of NHANES 2007-2018 data. Frontiers in nutrition 12, 1593174, doi:10.3389/fnut.2025.1593174 (2025).

16 Du, W. et al. The association between constipation and stroke based on the NHANES and Mendelian randomization study. Frontiers in neuroscience 17, 1276032, doi:10.3389/fnins.2023.1276032 (2023).

17 Tang, F. et al. Association between constipation and risk of stroke: a systematic review and meta-analysis. Frontiers in neurology 16, 1594535, doi:10.3389/fneur.2025.1594535 (2025).

18 Ananth, C. V. et al. Epidemiology and trends in stroke mortality in the USA, 1975-2019. International journal of epidemiology 52, 858–866, doi:10.1093/ije/dyac210 (2023).

19 Peh, A., O’Donnell, J. A., Broughton, B. R. S. & Marques, F. Z. Gut Microbiota and Their Metabolites in Stroke: A Double-Edged Sword. Stroke 53, 1788–1801, doi:10.1161/strokeaha.121.036800 (2022).

20 Zeng, J. et al. The mechanism of intestinal microbiota regulating immunity and inflammation in ischemic stroke and the role of natural botanical active ingredients in regulating intestinal microbiota: A review. Biomedicine & pharmacotherapy = Biomedecine & pharmacotherapie 157, 114026, doi:10.1016/j.biopha.2022.114026 (2023).

21 Tuz, A. A., Hasenberg, A., Hermann, D. M., Gunzer, M. & Singh, V. Ischemic stroke and concomitant gastrointestinal complications-a fatal combination for patient recovery. Frontiers in immunology 13, 1037330, doi:10.3389/fimmu.2022.1037330 (2022).

22 Xu, K. et al. Rapid gut dysbiosis induced by stroke exacerbates brain infarction in turn. Gut, doi:10.1136/gutjnl-2020-323263 (2021).

23 Lo, C. H. et al. Association of Ultra-processed Food and Unprocessed or Minimally Processed Food Consumption With Bowel Habits Among U.S. Adults. Clinical gastroenterology and hepatology : the official clinical practice journal of the American Gastroenterological Association 22, 2309–2318.e2305, doi:10.1016/j.cgh.2024.04.036 (2024).

24 Yun, Q. et al. Constipation preceding stroke, dementia and Parkinson’s disease in middle-aged and older adults: a population-based cohort study. Age and ageing 54, doi:10.1093/ageing/afaf257 (2025).

25 Sharma, A., Voigt, R. M., Goetz, C. G. & Keshavarzian, A. Parkinson Disease and the Gut: A Primer for Gastroenterologists. The American journal of gastroenterology, doi:10.14309/ajg.0000000000003508 (2025).

26 Wei, Y. H. et al. The gastrointestinal-brain-microbiota axis: a promising therapeutic target for ischemic stroke. Frontiers in immunology 14, 1141387, doi:10.3389/fimmu.2023.1141387 (2023).

